# Effect of combined cap-assisted, water-aided, and prone position colonoscopy for adenoma detection: A retrospective study

**DOI:** 10.1101/2020.10.23.20218263

**Authors:** Jihwan Ko, Byung Gu Ko, Seong Ho Han, Hyung Wook Kim

## Abstract

**Background:** The efficacy of cap-assisted, water-aided, and 12 o’clock-prone position colonoscopy as individual techniques for adenoma detection is well documented. However, the efficacy of the combination of the three colonoscopy methods is unclear. Therefore, the present study aimed to retrospectively compare the efficacy between combined-method colonoscopy (CMC) and standard colonoscopy (SC).

**Methods and Findings:** A total of 746 patients who underwent either CMC or SC, performed by two board-certified gastroenterologists between December 2019 and March 2020 at Baekyang Jeil Internal Medicine Clinic, were retrospectively evaluated. We evaluated polyp detection rate (PDR), adenoma detection rate (ADR), and mean number of adenomas detected per procedure (MAP). Statistical analysis for comparison between the groups was performed using the Student’s t-test, and ADR and PDR were analyzed using Fisher’s exact test. The study population was predominantly women (55.4%). The mean patient age (standard deviation) was 62.87 (±7.83) years. There was no significant difference in sex, number of fecal occult blood test-positive patients, and age between the two groups. The PDR, ADR, and proximal colon MAP were significantly higher in the CMC group than in the SC group (PDR: 59.8% vs. 84.9%, p < 0.001; ADR: 49.2% vs. 70.1%, p < 0.001; proximal colon MAP: 0.55 vs. 1.24, p < 0.001).

**Conclusions:** Compared with SC, CMC increases PDR, ADR, and MAP, especially proximal colon MAP. Therefore, CMC may be more useful than SC in clinical settings. This study is the first to evaluate the efficacy of the three techniques in combination.

## Introduction

Colorectal cancer is the third leading cause of new cancer cases and second leading cause of cancer deaths worldwide [1]. Colonoscopic screening of colorectal cancer has several advantages, such as high-sensitivity detection of cancer and precancerous lesions and single-step diagnosis and treatment [2]. Several studies have reported the efficacy of colonoscopy in preventing incident colorectal cancer and cancer deaths [3-6]. Although colonoscopic screening has many advantages, it has a single most important disadvantage: it is operator dependent for the detection of precancerous lesions and cancer [7,8]. To decrease this operator dependency, many quality parameters have been used, including adenoma detection rate (ADR) and mean number of adenomas detected per procedure (MAP) [9,10] Because these two parameters are important in improving the colonoscopic screening effect, many studies have been performed focusing on the improvement of cap-assisted colonoscopy [11], water-aided colonoscopy [12], and prone position colonoscopy [13]. Water infusion instead of air insufflation during the insertion phase increased ADR. Prone position reduces ileal intubation time. This is not directly related to the ADR, but there was a study that showed shorter insertion times to be associated with an increased rate of small colorectal adenomas [14].

We intended to search for a method for increasing ADR and MAP without using complicated devices that cannot be applied in the clinical setting [15] and decided to assess if there is any synergistic effect on application of all three methods at once. The aim of this study was to compare the ADR between combined-method colonoscopy (CMC) (clear cap-assisted, prone position, water-aided) and standard colonoscopy (SC) (left decubitus, air insufflating).

## Methods

### Study design

This single-center retrospective case-control study was conducted at Baekyang Jeil Internal Medicine Clinic, Busan, South Korea. This study was approved by the ethics committee of the Institutional Review Board (IRB) of Pusan National University Yangsan Hospital (IRB number: 05-2020-126) and conducted in accordance with the principles of the Declaration of Helsinki.

### Patients

Between December 2019 and March 2020, a total of 901 patients underwent colonoscopy at our hospital, and the procedures were performed by two board-certified gastroenterologists. In total, 442 patients underwent CMC, and 459 patients underwent SC. We analyzed the data for patients over 50 years of age. Of the 746 patients enrolled, 388 were included in the SC group and 358 in the CMC group. The only exclusion criterion was age below 50 years. The study groups were not randomized. Each patient was made to choose a doctor for their colonoscopy. In South Korea, the National Health Insurance System urges people over the age of 50 years to take a fecal occult blood test(FOBT) annually; when the test is positive, it provides financial support for undergoing colonoscopy. The proportion of FOBT-positive patients could affect ADR [16]; hence, we compared the number of FOBT-positive patients in each group. Fortunately, age, sex, and number of positive FOBT were not significantly different (Table 1).

**Table 1.**
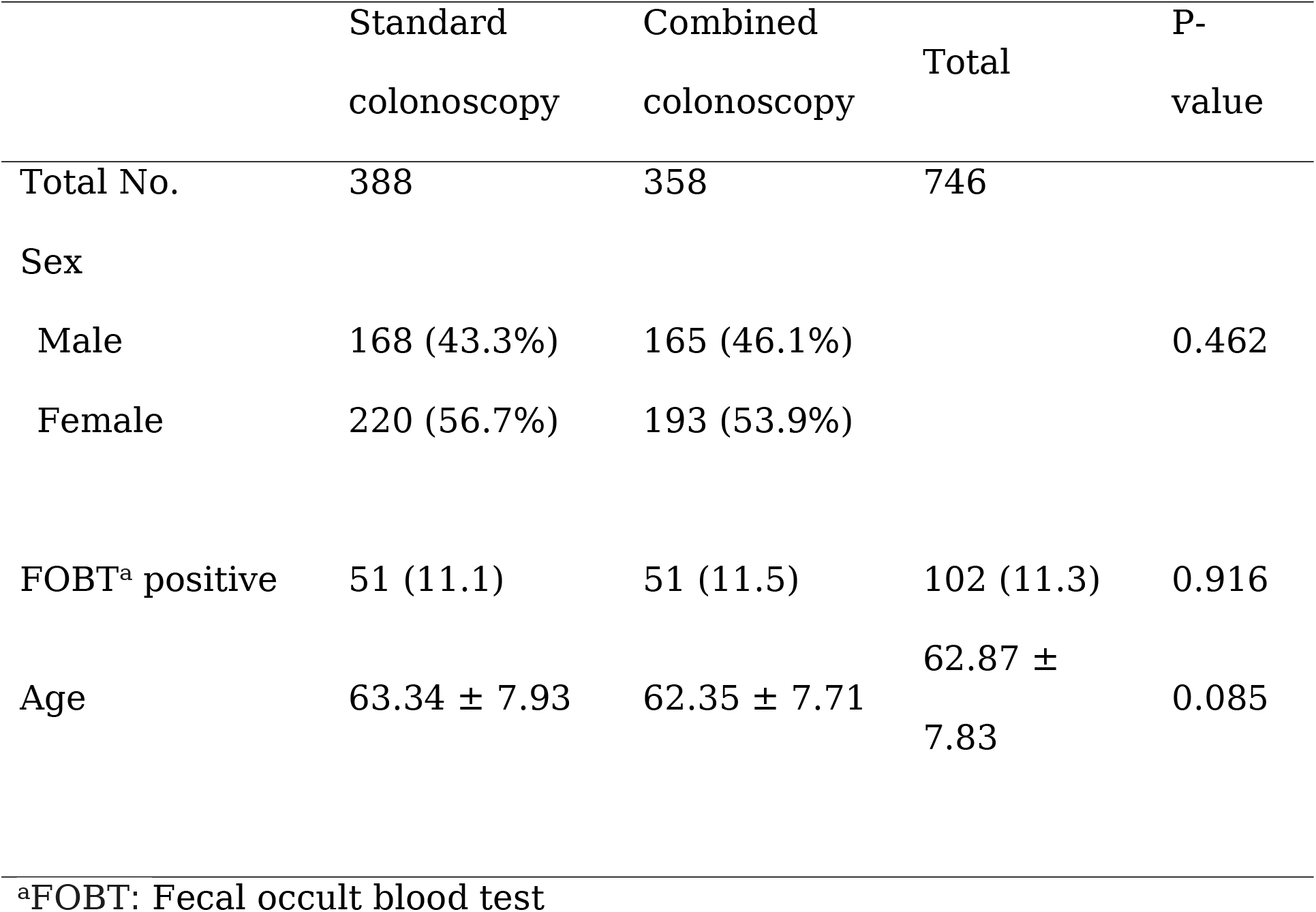
Baseline data and clinical indications for colonoscopy (n = 746)

|  | Standard<br>colonoscopy | Combined<br>colonoscopy | Total | P-value |
| --- | --- | --- | --- | --- |
| Total No. | 388 | 358 | 746 |  |
| Sex |  |  |  |  |
| Male | 168 (43.3%) | 165 (46.1%) |  | 0.462 |
| Female | 220 (56.7%) | 193 (53.9%) |  |  |
| FOBT<sup>a</sup> positive | 51 (11.1) | 51 (11.5) | 102 (11.3) | 0.916 |
| Age | 63.34 ± 7.93 | 62.35 ± 7.71 | 62.87 ±<br>7.83 | 0.085 |

<sup>a</sup>FOBT: Fecal occult blood test

### Ethical approval

All procedures performed in studies involving human participants were performed in accordance with the ethical standards of the institutional and/or national research committee and with the 1964 Helsinki Declaration and its later amendments or comparable ethical standards. Informed consent was obtained from all individual participants included in the study.

### Endoscopic procedure

In the present study, two board-certified gastroenterologists performed colonoscopies, among which one had performed more than 10,000 examinations before the study period with SC and the other had performed more than 3,000 examinations before the study period with CMC. Because there was no association between procedural volume and ADR, we assumed that procedural volume difference between the two doctors should not affect ADR [17]. Colonoscopies were performed after bowel preparation with 2 L polyethylene glycol plus ascorbic acid solution (Coolprep, Taejun, Seoul, Korea; Readyfree, Intropharm Tech., Gyeonggi-do, Korea). Colonoscopies were performed with EPK-i5000 (Pentax EPKi processor) and EC38-i10F colonoscopes (Pentax, Tokyo, Japan). A transparent cap was attached to the tip of the colonoscope (Finemedix Co. Ltd, Daegu, Korea). All examinations were performed under conscious midazolam-induced sedation. Endoscopic observation and therapeutic intervention for detected polyps were performed during the withdrawal phase. Both the groups started in the left decubitus position, but as soon as the colonoscope reached the sigmoid colon, the CMC group changed to the 12 o’clock-prone position and kept in this position until the end of the procedure. The CMC group used the Pentax endoscope’s own waterjet system for visualizing the colon pathway, and the SC group used air for visualizing the colon pathway.

### Polyps

Polyps with a diameter ≥ 5 mm were removed by endoscopic mucosal resection or cold snare polypectomy. Polyps with a diameter <5 mm were removed with biopsy forceps. All removed polyps were pathologically examined, and the number of adenomas was determined. In this study, we defined polyp detection rate (PDR) as the proportion of patients with at least one polyp and ADR as the proportion of patients with at least one adenoma.

### Outcome measures and statistical analysis

The principal outcome was the comparison of ADR between SC and CMC groups. The secondary outcome was the comparison of PDR and MAP. Proximal and distal colon MAP was also evaluated.

Statistical comparisons between the two groups were performed using the Student t-test, and ADR and PDR were analyzed using Fisher’s exact test. P < 0.05 was considered to indicate statistical significance. Statistical analyses were performed using SPSS (version 12, IBM Corp., Armonk, NY, USA).

## Results

### Patient characteristics

Baseline characteristics were not significantly different between the two groups (Table 1). A total of 746 colonoscopies were evaluated. The study population was composed predominantly of women (55.4%), and the mean age ± SD was 62.87 ± 7.83 years. The indications for colonoscopy were categorized into the following: first, FOBT-positive, financially supported by the National Health Insurance System; and second, FOBT negative or not undergoing the FOBT test, paying their own money for evaluating the presence of polyps or cancer. There was no significant difference in sex or study indication and age between the two groups.

### Polyp detection rate

A total of 536 people (71.8%) had >1 polyp: 232 (PDR: 59.8%) in the SC group and 304 (PDR: 84.9%) in the CMC group. The PDR was significantly higher in the CMC group (p < 0.001).

### Adenoma detection rate

A total of 442 people (59.2%) had >1 adenoma: 191 (ADR: 49.2%) in the SC group and 251 (70.1%) in the CMC group. The ADR was significantly higher in the CMC group (p < 0.001), and MAP was also significantly higher in the CMC group than that in the SC group (1.69 ± 1.93 vs. 1.06 ± 1.59, respectively; p < 0.001). Proximal MAP significantly increased than did distal MAP (Table 2).

**Table 2.**
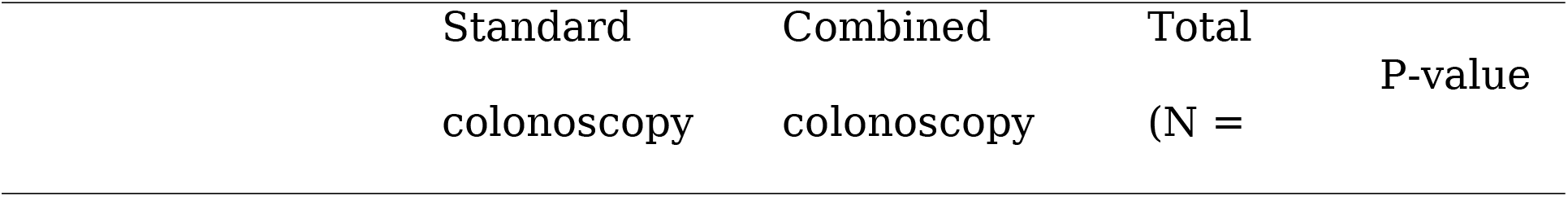
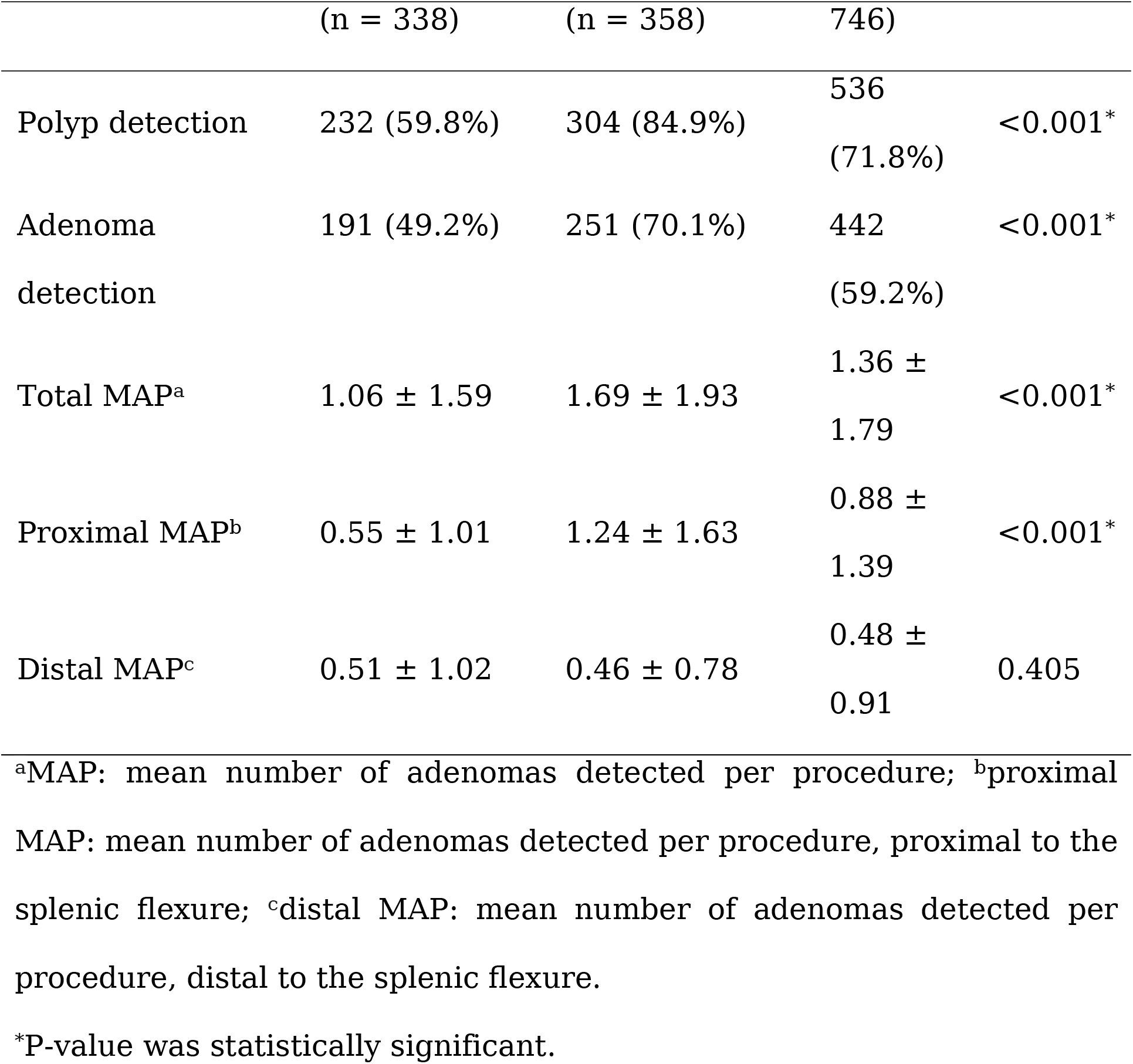
Quality indicators by colonoscopy methods

## Discussion

Colonoscopic screening is widely used for preventing colorectal cancer. The two important quality indicators of colonoscopy are ADR and MAP [9,10]. Several methods, such as Endocuff, Endocuff Vision, or EndoRings, have been studied for increasing the two quality indicators [15]. However, in countries with a National Health Insurance System, private clinics have limitations for adopting these methods. We, therefore, decided to determine whether there is a possibility to increase the ADR by combining several simple methods, such as water-aided, prone position, and clear cap-assisted colonoscopy. In this study, we found that the PDR, ADR, MAP, and proximal MAP were significantly higher in the CMC group than those in the SC group. Several studies reported that each of the cap-assisted colonoscopy [11], water-aided colonoscopy [12], and prone 12 o’clock-position [13,14] could improve ADR compared to SC. However, to the best of our knowledge, this is the first study that achieved an ADR >70% in the general population. This study suggests that an endoscopist can detect more adenomas, especially in the proximal colon, because the combined method enables fewer polyps to be missed. To the best of our knowledge, no previous study has assessed the efficacy of CMC.

This study has some limitations. First, this was a single-center retrospective study with a small number of subjects. Second, because of the limitation of the retrospective study design, we could not investigate the patients’ family history of colorectal cancer and body mass index [18,19]. Third, each doctor’s study time could not be evaluated, thereby introducing potential selection bias.

In conclusion, ADR and MAP of CMC were significantly higher than those of SC, suggesting that CMC may be more useful than SC. Therefore, further multicenter randomized controlled trials on each combinable method should be conducted to assess possible synergistic effects.

## Data Availability

All data generated or analysed during this study are included in this published article.

## Abbreviations

ADR: Adenoma detection rate
CMC: Combined method colonoscopy
FOBT: Fecal occult blood test
IRB: Institutional review board
MAP: Mean number of adenomas detected per procedure
PDR: Polyp detection rate
SC: Standard colonoscopy

## Acknowledgments

We would like to thank Editage (www.editage.co.kr) for editing and reviewing this manuscript for English language.

